# The utility of established prognostic scores in COVID-19 hospital admissions: a multi-centre prospective evaluation of CURB-65, NEWS2, and qSOFA

**DOI:** 10.1101/2020.07.15.20154815

**Authors:** Patrick Bradley, Freddy Frost, Kukatharmini Tharmaratnam, NW-CORR Collaborators, Dan Gower Wootton

## Abstract

**Introduction:** The COVID-19 pandemic is ongoing yet, due to the lack of a COVID-19 specific tool, clinicians must use pre-existing illness severity scores for initial prognostication. However, the validity of such scores in COVID-19 is unknown.

**Methods:** The North West Collaborative Organization for Respiratory Research (NW-CORR) performed a multi-centre prospective evaluation of adult patients admitted to hospital with confirmed COVID-19 during a two-week period in April 2020. Clinical variables measured as part of usual care at presentation to hospital were recorded, including the CURB-65, NEWS2, and qSOFA scores. The primary outcome of interest was 30-day mortality.

**Results:** Data were collected for 830 people with COVID-19 admitted across 7 hospitals. By 30 days, a total of 300 (36.1%) had died and 142 (17.1%) had been in ICU. All scores underestimated mortality compared to their original validation in non-COVID-19 populations, and overall prognostic performance was generally poor. Among the ‘low risk’ categories (CURB-65<2, NEWS2<5, qSOFA<2) 30-day mortality was 16.7%, 32.9% and 21.4%, respectively. Multivariable logistic regression identified features of respiratory compromise rather than circulatory collapse as most relevant prognostic variables.

**Conclusion:** All existing prognostic scores evaluated here underestimated adverse outcomes and performed sub-optimally in the COVID-19 setting. New prognostic tools including a focus on features of respiratory compromise rather than circulatory collapse are needed. We provide a baseline set of variables which are relevant to COVID-19 outcomes and may be used as a basis for developing a bespoke COVID-19 prognostication tool.

**Key Messages:**

- **What is the key question?** Do well-established illness severity scores have prognostic value in COVID-19?
- **What is the bottom line?** All scores appeared to underestimate mortality in COVID-19 and prognostic performance was generally poor, and importantly could not discriminate those patients at very low risk of death within 30 days.
- **Why read on?** In this multi-centre prospective evaluation of CURB-65, NEWS2 and qSOFA we comprehensively evaluate score performance and also identify variables which may be of use in COVID-19 prognostication.

## Introduction

The novel coronavirus SARS-CoV-2 is causing a global pandemic of the infectious disease termed COVID-19. COVID-19 is frequently associated with a pneumonia syndrome and the large ISARIC observational study estimates a case fatality rate of 33% among those admitted to hospital [1]. Prognostic scores can improve clinical decision making and pre-COVID-19 several scores had been extensively validated and supported by national and international guidelines for application in the context of acute infectious disease [2–4].

The CURB-65 score (confusion, urea, respiratory rate, blood pressure and age above or below 65 years) is a community acquired pneumonia (CAP) specific tool for predicting all-cause mortality within 30 days. CURB-65 has been validated across large, diverse patient populations and has been endorsed by national and international guidelines as an aid to clinical decision making [5–9]. The National Early Warning Score 2 (NEWS2) is a scoring system based upon routine physiological measurements and its implementation into all English National Health Service (NHS) hospitals had been mandated in the pre-COVID-19 era [10]. NEWS2 is a disease agnostic early warning tool used to trigger escalation of care in the deteriorating patient, with high scores being associated with death or unanticipated ICU admission within 24 hours [2]. The quick Sequential [Sepsis-related] Organ Failure Assessment (qSOFA) score is a tool for predicting mortality and ICU admission among patients with suspected infection in pre-hospital, emergency department and ward settings. It has been validated through large datasets, and has gained prominence following its recommendation by the Sepsis-3 task force [11,12].

At the onset of the UK epidemic, in the absence of COVID-19 specific prognostic tools, CURB-65, NEWS2 and qSOFA and remained in widespread use but little was known about their validity in the COVID-19 setting. The aim of this study was to determine the performance characteristics of these scores in the context of COVID-19 and to investigate potential components of a COVID-19 specific prognostication tool for future validation.

## Methods

### Study setting and participants

The North West Collaborative Organization for Respiratory Research (NW-CORR) collected data during the two-week period 1/4/2020 to 14/4/2020 on prospective, adult, COVID-19 admissions at 7 acute hospitals in North West England. NW-CORR constitutes a group of research interested, respiratory, specialist trainee grade doctors and the recruiting centres were those with an NW-CORR member available. Collaborators were asked to record routinely collected clinical data for consecutive patients admitted to their hospitals who met the Public Health England inpatient case-definition for COVID-19 [13], and had a positive SARS-CoV-2 polymerase chain reaction (PCR) test. There were no exclusion criteria. Institutional approval for data collection and anonymised data collation was obtained from all sites and were a combination of Caldicott guardian and research/audit committee approvals. No approach to the patient was made and only fully anonymised routinely available clinical information was collated; on this basis consent was not required under guidance from the NHS Human Research Authority [14].

### Outcomes and prognostic scores

Data collected included demographic characteristics, vital signs and blood test results that were recorded at the point of presentation to hospital, including the constituent components of the CURB-65, NEWS2 and qSOFA scores. The variables measured for each of these were recorded (see Table 1). At the point of data entry, collaborators were also asked to comment on the presence or absence of consolidation on chest radiography. Outcomes including all-cause mortality, intensive care unit (ICU) admission and discharge were recorded following a 30-day period from each patient’s admission. ‘Early death’ was classified as death within 72 hours of admission.

**Table 1:** Components of the NEWS2, qSOFA and CURB-65 scores

| CURB-65 | NEWS2 | qSOFA |
| --- | --- | --- |
| • Confusion<br>• Urea >7 mmol/L<br>• Respiratory rate 30/min<br>• Blood pressure: systolic <90 or diastolic 60 mmHg<br>• Age 65 years | • Respiratory rate<br>• SpO<sub>2</sub><br>• Supplemental O<sub>2</sub> use<br>• Heart rate<br>• Altered consciousness<br>• Temperature | • Altered mental status<br>• Respiratory rate 22/min<br>• Blood pressure: systolic 100 mmHg |

### Data handling

Anonymised study data were collated centrally and managed using the secure, web-based software platform REDCap (Research Electronic Data Capture, Vanderbilt University, USA) hosted at the University of Liverpool.

### Statistical analysis

Each score was assessed individually against their validated outcomes and overall for their ability to identify people at risk of mortality within 72 hours (early death) and 30 days of admission. This analysis included sensitivity and specificity of each score’s respective risk strata followed by an evaluation of discrimination and calibration in keeping with TRIPOD guidelines [15]. Discriminatory ability was assessed by comparison of the corresponding receiver operator characteristics (ROC) curves with computation of area under curve (AUC). Calibration was assessed visually by plotting the observed risk for a score’s individual strata against published reference risk derived from their original validation. In order to allow direct comparison of the clinical scoring systems, only patients with complete data for all variables were included in comparative statistical analyses.

Multiple logistic regression models were fitted for each of the outcome variables (30 days mortality, 72 hours mortality and ICU admission) using each score (CURB-65, NEWS2 and qSOFA). With the aim of identifying variables relevant to COVID-19 outcomes and in order to assess the association of each individual variable (e.g. age, respiratory rate, etc.) with each of the outcomes, multiple regression modelling using all variables was fitted by applying backward variable selection. Data heterogeneity introduced by differences among hospitals was assessed by adding a random intercept in the model. However, clustering by hospital did not improve the accuracy of the model and the final models did not include a random term. The performance of the fitted models was assessed by sensitivity, specificity, positive predictive value (PPV), negative predictive value (NPV) and area under curve (AUC). These analyses performed on patients with complete data using statistical software R version 3.5.3 with internal cross validation based on bootstrap sampling method.

## Results

Data were collected and recorded for 830 patients admitted to 7 hospitals in the north-west of England, encompassing both secondary and tertiary care hospitals, see Supplement. Clinical characteristics and observations at admission are presented in Table 2. In general, there was minimal missing data with >99% completeness for all constituent variables of the three prognostic scores, Table 2. Overall, 509/830 (61.3%) were male with a median age of 70 years [inter-quartile range [IQR] 58 to 80] and Rockwood clinical frailty score (CFS) 4 [IQR 2 to 6]. Within 72 hours of admission, 63/830 (7.6%) patients had died and 125/830 (15.0%) had been admitted to ICU. At 30 days, 300/830 (36.1%) had died, 452/830 (54.5%) has been discharged and 78/830 (9.4%) remained in hospital. During the 30-day period, 142 (17.1%) were admitted to critical care, of whom 65 (45.8%) died. A comparison of clinical characteristics based on 30-day and 72-hour mortality is presented in Table 2 and Table S1 respectively.

**Table 2:** Baseline demographics and clinical characteristics with comparison by outcome at 30 days. Data are presented as n(%) or median (IQR). P-values are calculated using Fisher’s exact test for categorical variables, Mann-Whitney (M) test for continuous variables since none of the continuous variables follow the assumption of normality (Shapiro normality test p-value=<0.001 for all of them).

|  | Missing | All | Death by 30 days |  |  |
| --- | --- | --- | --- | --- | --- |
|  |  |  | Alive | Dead | p |
| n |  | 830 | 530 | 300 |  |
| **Characteristics at presentation** |  |  |  |  |  |
| Age (Years) | 1 (0.1%) | 70 (58,80) | 65 (55,76) | 76 (67,85) | <0.001 |
| Sex (Male) | 0 (0.0%) | 509 (61.3%) | 309 (58.3%) | 200 (66.7%) | 0.02 |
| CFS | 81 (9.8%) | 4 (2,6) | 3 (2,5) | 5 (3,6) | <0.001 |
| Temp (°C) | 3 (0.4%) | 37.5 (36.8,38.2) | 37.4 (36.8,38.1) | 37.6 (36.8,38.4) | 0.15 |
| Respiratory rate (/min) | 2 (0.2%) | 24 (20,28) | 22 (20,26) | 24 (21,30) | <0.001 |
| Heart rate (bpm) | 1 (0.1%) | 94 (82,108) | 95 (82,108) | 91 (80,110) | 0.28 |
| Systolic BP (mmHg) | 2 (0.2%) | 130 (115,145) | 130 (117,145) | 131 (113,145) | 0.78 |
| Diastolic BP (mmHg) | 2 (0.2%) | 75.0 (65.0,84.0) | 76.0 (67.0,85.0) | 72 (63,81) | <0.001 |
| SpO2 (%) | 1 (0.1%) | 94 (90,96) | 94 (91,96) | 93 (89,96) | 0.002 |
| FiO2 (%) | 2 (0.2%) | 21 (21,32) | 21 (21,28) | 24 (21,50) | <0.001 |
| SpO2/FiO2 ratio | 2(0.2%) | 414 (283, 448) | 429 (339, 452) | 354 (189, 443) | <0.001 |
| Confusion | 0 (0.0%) | 183 (22.0%) | 90 (17.0%) | 93 (31.0%) | <0.001 |
| **Investigations** |  |  |  |  |  |
| Urea (mmol/L) | 8 (1.0%) | 7.2(4.9,11.1) | 6.3(4.6,9.2) | 9.8(7.0,15.1) | <0.001 |
| Consolidation | 4 (0.5%) | 684 (82.8%) | 425 (80.5%) | 259 (86.9%) | 0.02 |
| CRP (mg/L) | 10 (1.2%) | 104.0(47.8,173.1) | 92.0(42.5,158.0) | 117.0(59.0,195.0) | <0.001 |
| WCC (10<sup>9</sup>/L) | 4 (0.5%) | 7.4(5.6,10.1) | 7.3(5.5,9.8) | 7.6(5.7,10.6) | 0.20 |
| Neutrophils (10<sup>9</sup>/L) | 7 (0.8%) | 5.8(3.9,8.55) | 5.6(3.8,7.9) | 5.9(4.0,9.1) | 0.26 |
| Lymphocytes (10<sup>9</sup>/L) | 9 (1.1%) | 0.8(0.6,1.1) | 0.9(0.6,1.2) | 0.7(0.5,1.0) | <0.001 |
| NLR | 9 (1.1%) | 154.0(91.0,201.0) | 155.0(95.5,201.0) | 152.5(85.3,201.0) | 0.62 |

**Abbreviations:** CFS=Clinical frailty scale; BP=blood pressure; SpO2=oxygen saturation by pulse oximetry; FiO2=fraction of inspired oxygen; CRP=C-reactive protein; WCC=White cell count; NLR=Neutrophil/Lymphocyte ratio

The discriminatory ability of each score was assessed for death within 30 days, death within 72 hours, and admission to critical care, and are presented in Figure 2. In general, performance was modest, with AUCs ranging from 0.62 to 0.77. Calibration was computed by comparing predicted risk from each score against the respective observed risk in the study cohort. Visual comparison each calibration plot confirmed slopes >1 and intercepts >0 suggestive of underestimation, see Figure 3.

**Figure 1:**
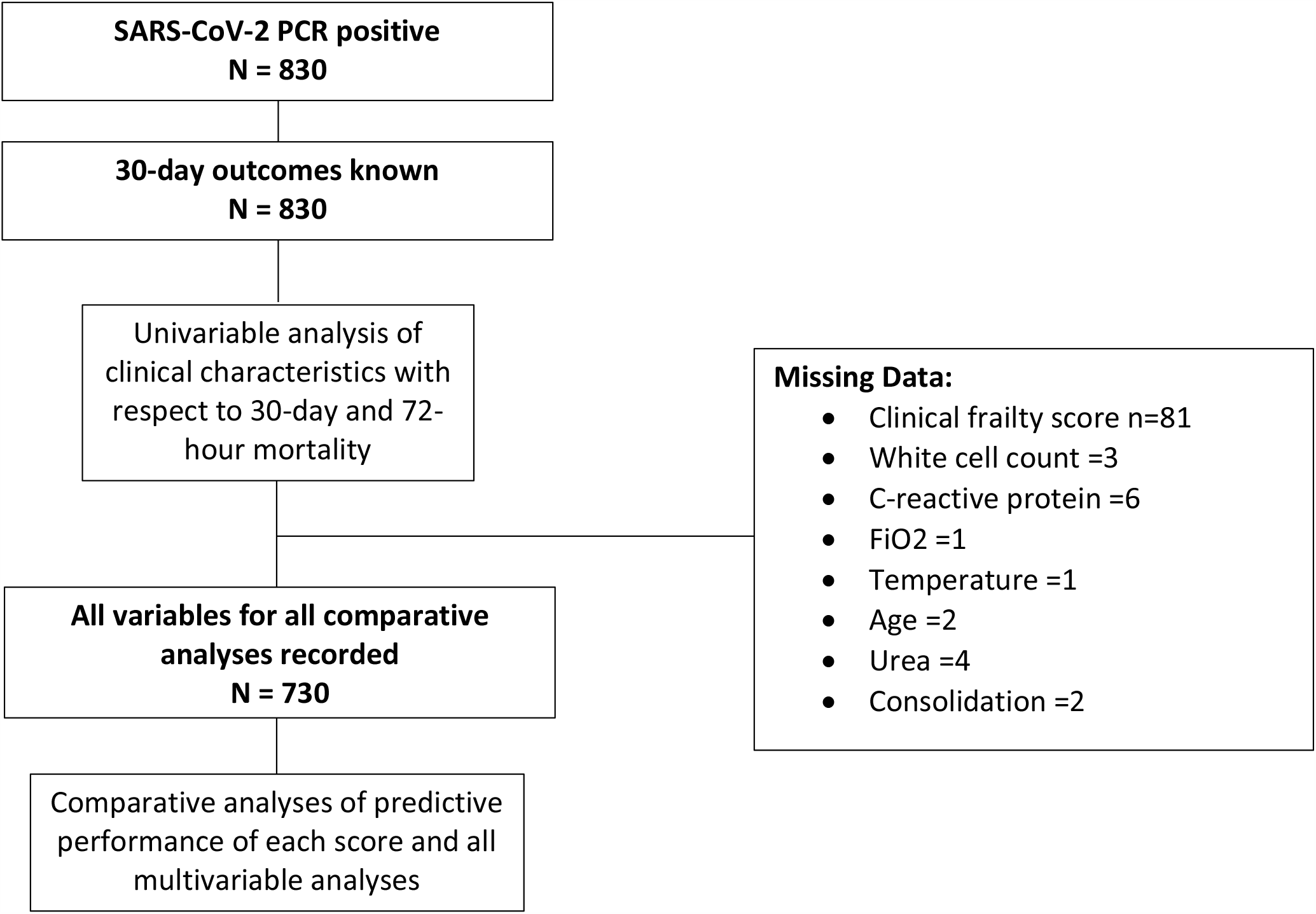
Patient flow chart describing cohort for each analysis and missing data.

**Figure 2:**
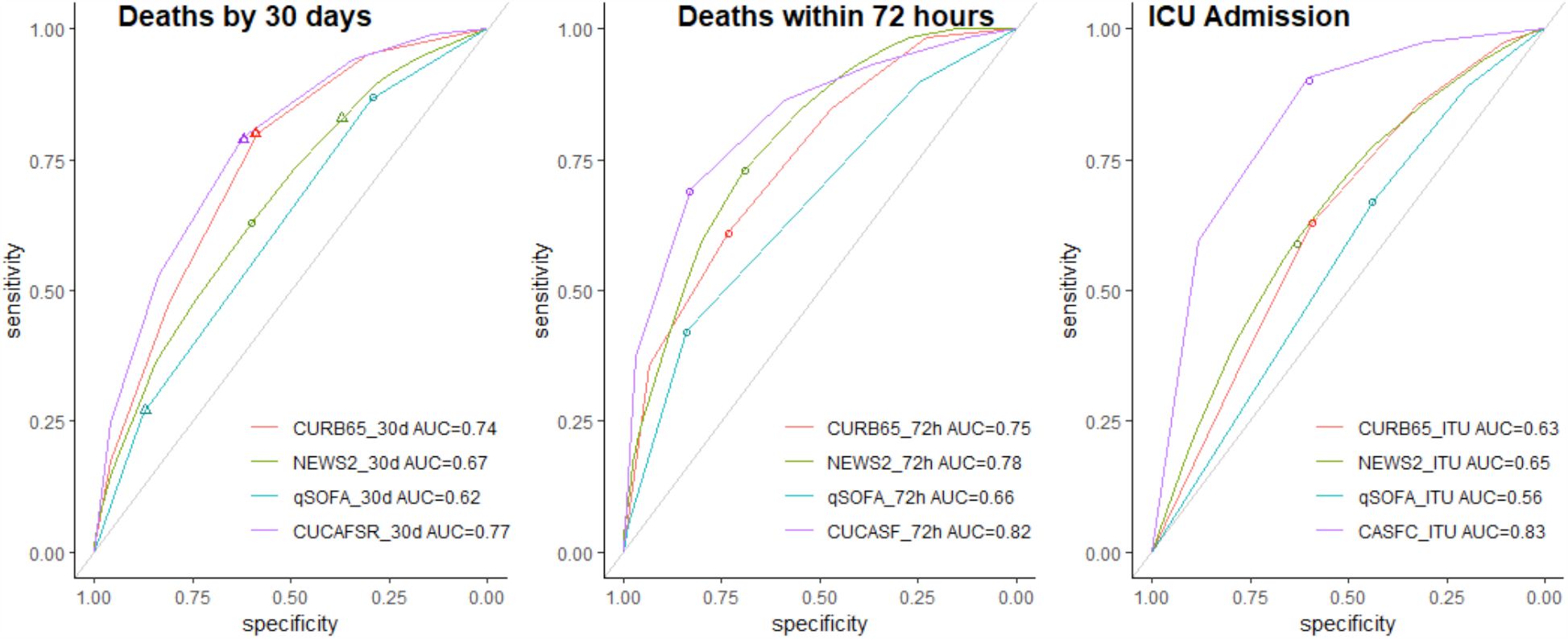
ROC plots for Death by 30 days based on the models defined as the sum of the corresponding predictors. Triangles denote the points with the sensitivity and specificity achieved (cut-offs 2, 5, 2 and 4 for tools CURB65, NEWS2, qSOFA and CUCAF-SR, respectively). Circles denote the sensitivity and specificity achieved by the optimal threshold from fitted models

**Figure 3:**
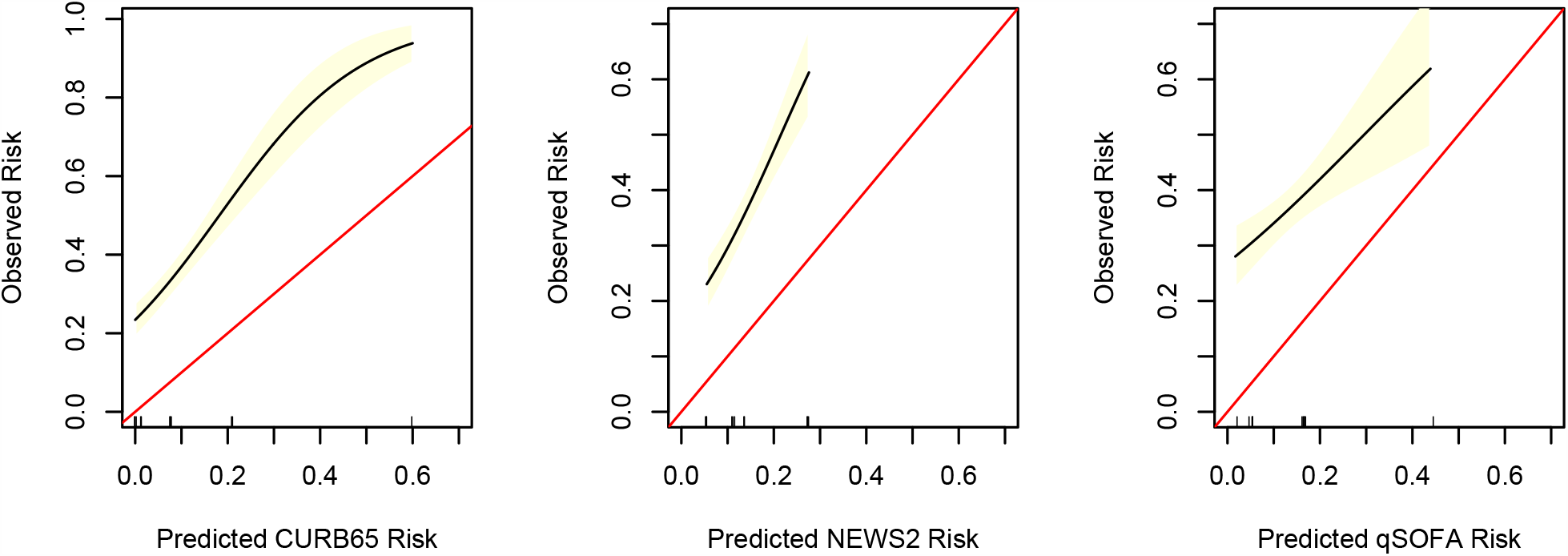
Calibration plots of the predicted risk of 30-day mortality (based upon published validation studies) for CURB65 [16], NEWS2 [10] and qSOFA [17] against observed risk in COVID-19 hospital admissions.

To test scores’ performance at their individual validated thresholds, the sensitivity, specificity, negative predictive value and positive predictive value were calculated and are presented in Table 3. Overall, for 30-day mortality, scores failed to accurately identify a low risk group, with mortality in the lowest risk strata ranging from 16% to 33. For 72-hour mortality, a CURB-65 threshold of 2 and NEWS2 threshold of 5 both identified a low-risk group with just 2% mortality and NEWS2 achieved sensitivity of 92% with NPV of 98%. All scores performed poorly in predicting admission to ICU, see Table S2. Relative likelihood of mortality at each stratum of each score is presented in Table 4.

**Table 3:**
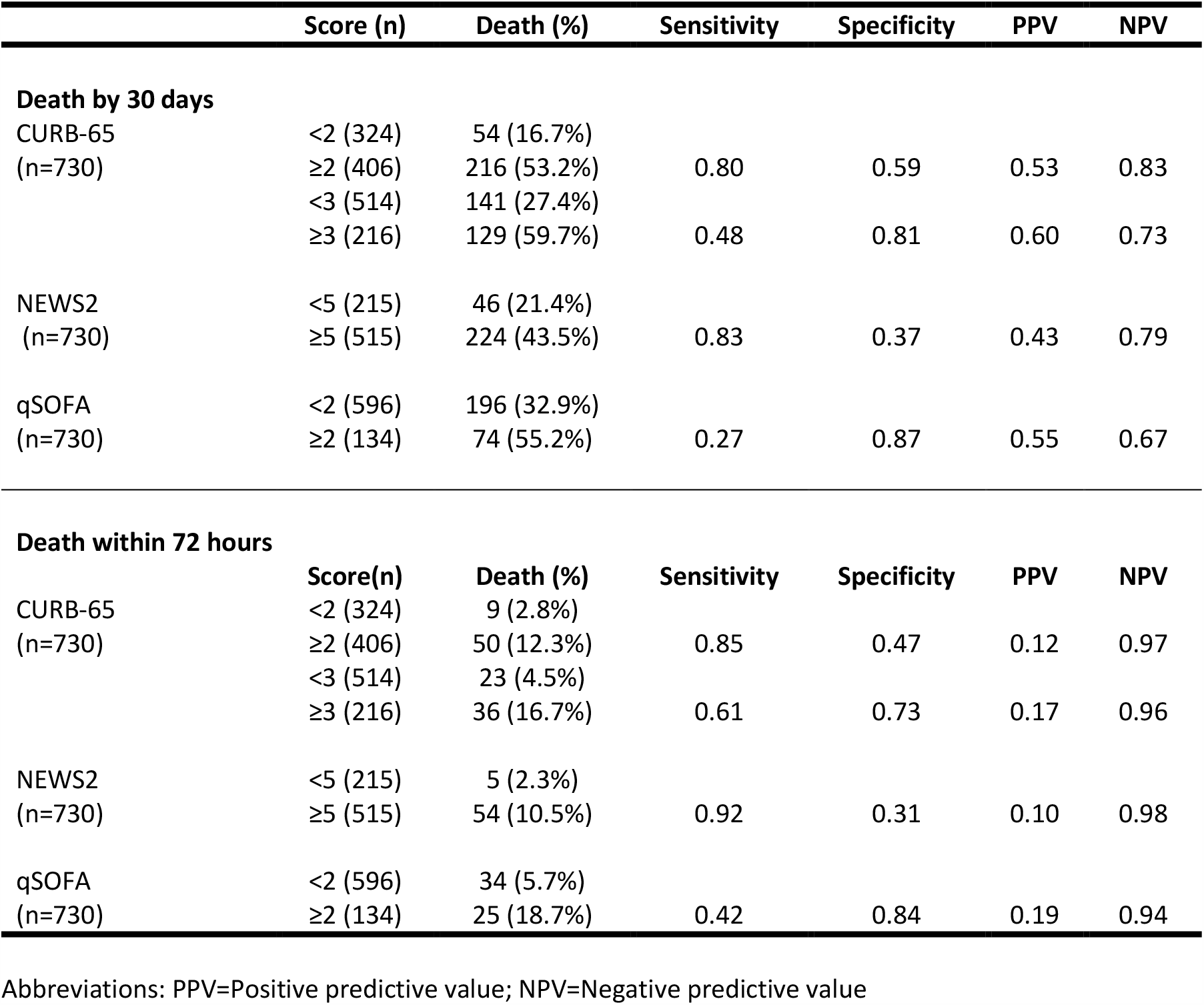
Diagnostic performance of individual scores for 30-day and 72-hour mortality

**Table 4:** Odds ratio of death within 30 days and 72 hours for the individual strata of each score

| **30-day mortality** |  |  |  |
| --- | --- | --- | --- |
| **Model** | **Score** | **OR (95%CI)** | **p-value** |
| **CURB-65** | 0 (reference) |  |  |
|  | 1 | 3.40 (1.74 – 6.62) | <0.001* |
|  | 2 | 9.10 (4.82 – 17.18) | <0.001* |
|  | 3 | 12.99 (6.76 – 24.95) | <0.001* |
|  | ≥4 | 26.64 (12.22 – 56.06) | <0.001* |
| **NEWS2** | <5 (reference) |  |  |
|  | ≥5 | 2.83 (1.95 - 4.09) | <0.001* |
| **qSOFA** | <2 (reference) |  |  |
|  | ≥2 | 2.52 (1.72 - 3.68) | <0.001* |
| **72h mortality** |  |  |  |
| **Model** | **Score** | **OR (95%CI)** | **p-value** |
| **CURB-65** | 0 (reference) |  |  |
|  | 1 | 7.46 (0.92 – 60.35) | 0.060 |
|  | 2 | 12.09 (1.57 – 93.02) | 0.017* |
|  | 3 | 16.89 (2.20 – 129.56) | 0.007* |
|  | ≥4 | 70.93 (9.29 – 541.79) | <0.001* |
| **NEWS2** | <5 (reference) |  |  |
|  | ≥5 | 4.92 (1.94 - 12.48) | 0.001* |
| **qSOFA** | <2 (reference) |  |  |
|  | ≥2 | 3.79 (2.18 - 6.61) | <0.001* |

**Table 5:**
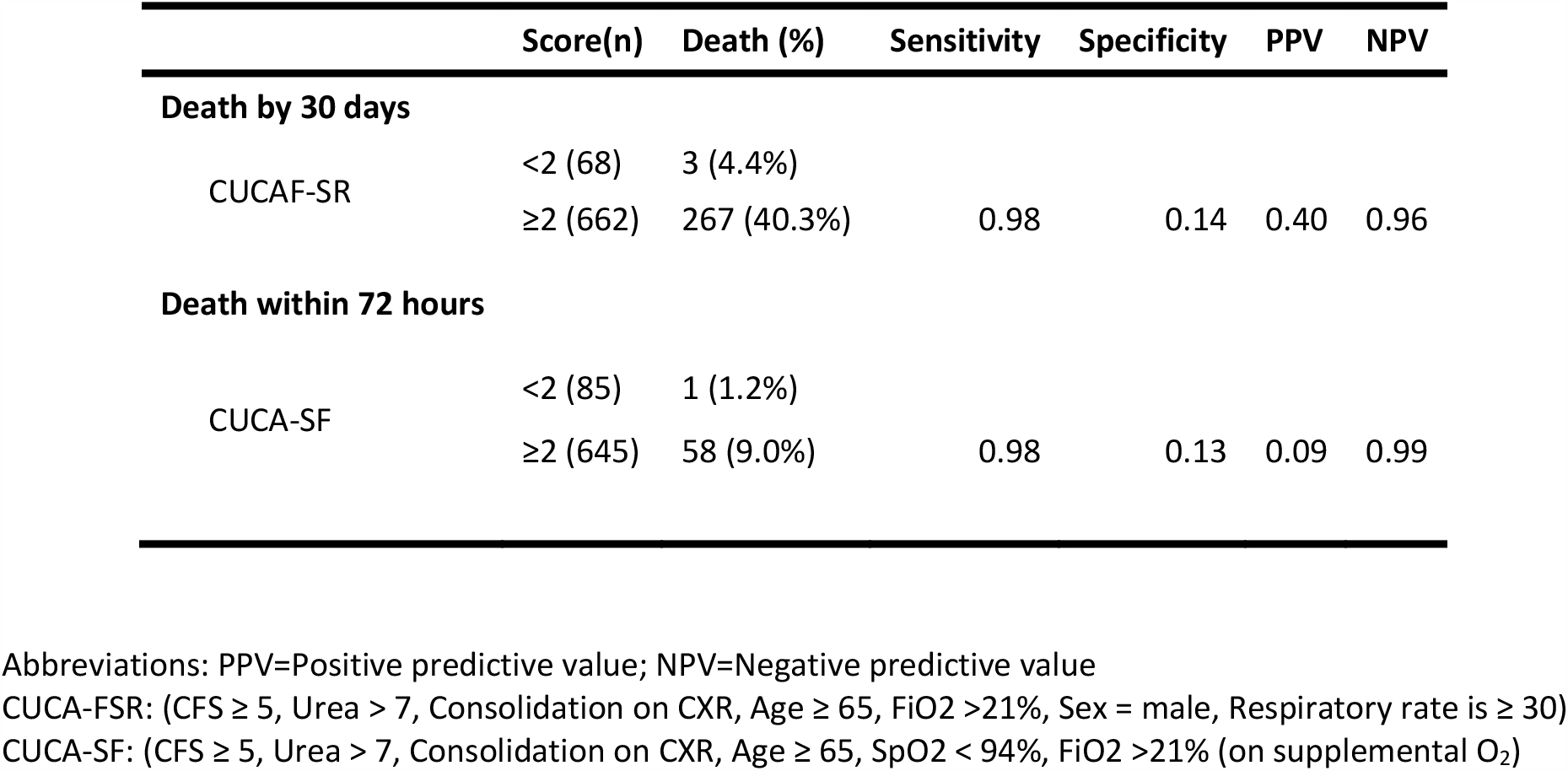
Diagnostic performance of logistic regression models fitted for 30-day mortality and 72-hour mortality

When all individual variables were considered, multivariable logistic regression revealed that confusion and blood pressure (BP) were less relevant to 30-day mortality than urea, respiratory rate and age when computed as part of the CURB-65 score, see Table S3. In a similar fashion, the most severe BP sand HR strata in NEWS2 (score of 3: systolic BP ≤90mmHg, HR>130 or <41/min) were not independently associated with poorer outcomes in the NEWS2 model, see Table S3, whereas the corresponding fraction of inspired oxygen (FiO2) and respiratory rate strata were relevant to mortality (OR 2.00 [1.4 to 2.8], p<0.001 and OR 1.9 [1.3 to 2.9], p=0.003 respectively).

Finally, a backwards selection multivariable model fitted for each outcome identified *de novo* a set of variables independently associated with 30-day mortality (CFS, Urea, Consolidation, Age, FiO2, Sex, Respiratory rate, referred to herein as CUCAF-SR) and a similar set of variables for 72-hour mortality (CFS, Urea, Consolidation, Age, SpO2, FiO2, referred to herein as CUCA-SF). When compared to the existing scores, CUCA-SF was superior to NEWS2 and CURB-65 when predicting early mortality (AUC 0.82 vs 0.61 and 0.75 respectively), whereas the CUCAF-SR model was similar to CURB-65 when predicting 30-day mortality (AUC 0.77 vs 0.74), see Figure 2. However, mortality in those with a ‘low risk’ CUCAF-SR score of 0 or 1 (maximum = 7), remained high at 4.4%. For ICU admission, CAS-FC (CFS+Age+SpO2+FiO2+CRP), outperformed existing prognostic scores, see Suppl S4.

## Discussion

We analysed the accuracy with which admission CURB-65, NEWS2 and qSOFA scores predict early mortality, late mortality and ICU admission in the context of COVID-19. In general, calibration was poor as all three scores underestimated the risk of adverse outcomes. CURB-65 and qSOFA both performed poorly in comparison to their respective standard applications, suggesting their utility is limited in COVID-19. In contrast, NEWS2 and CURB-65 were better at predicting early death, defined here as death <72 hours, where a NEWS2 threshold of 5 (the recommended threshold for urgent intervention) showed excellent negative predictive value. We went on to derive two sets of parameters which, when combined on admission with COVID-19, may provide a more accurate prediction of mortality and may provide a useful basis for predictive scores to be validated in larger datasets.

CURB-65 was designed and validated as a tool for helping healthcare workers decide which patients with community acquired pneumonia (CAP) could be safely managed in the community. A score of <2 is consistently associated with low mortality rates of 0.4-2.8% at 30-days [6,16]. However here, in the COVID-19 setting, a CURB-65 score of <2 was associated with a mortality of 17%. In the pre-COVID era, at the time of presentation to hospital with CAP, it was extremely unusual for the medical team to know the causal pathogen and although it was well recognised that there was a range of virulence among the possible viral and bacterial causes of CAP, this data confirms that SARS-CoV-2 is a highly virulent outlier. This finding has particular relevance to the evolving pandemic since, as transmission reduces, SARS-CoV-2 will become one of numerous endemic causes of CAP in many countries. It will therefore be important to recognise that this will reduce the performance of CURB-65 on undifferentiated CAP cases and makes a strong case for the implementation of rapid diagnostics to determine the aetiology of CAP. Whilst these data suggest low CURB-65 scores may not support early COVID-19 discharge, higher scores may still have value in predicting particularly poor outcomes given CURB-65 scores ≥3 were associated with death in 60% of cases, compared to just 22% in the pre-COVID era [5]. On that basis, high scores could prompt early escalation planning and inform discussions with patients and their families.

NEWS2 has been widely implemented in English hospitals as simple score consisting of routine physiological measurements. Whilst it is most widely recognised as a simple tool to identify inpatients in need of urgent or emergent medical attention based on changing physiological measurements, it is also often utilised in the emergency department where it has been validated in a number of syndromes including sepsis and acute dyspnoea [17,18]. We found that a NEWS2 score <5 accurately identified a group of patients at low risk group for early mortality, however it was less successful when 30-day mortality was considered. Our findings are based on a single measure of NEWS2 on admission and future longitudinal work would be needed to confirm if established NEWS2 trigger thresholds remain valid for inpatients with COVID-19.

qSOFA was validated for use amongst hospital inpatients and emergency department admissions as a simple and accurate way to identify people with infections at higher risk of poor outcomes [11,12,19]. Early data from China suggested those who survived COVID-19 had lower qSOFA scores, a finding replicated here [20]. However, in our study, the median qSOFA in those who died within 30 days was <2 and mortality in this ‘low risk’ qSOFA group was 32.5%. Taken together, the poor overall discriminatory ability of qSOFA and the poor diagnostic performance seen here suggests a qSOFA score on admission is not a useful prognostic tool in COVID-19. These findings are supported by a recent study which found qSOFA was low in people with COVID-19 admitted to critical care and could not reliably identify those at risk of death [21].

The poor performance of qSOFA is interesting given it was derived from cohorts of patients with sepsis; a syndrome defined as “life-threatening organ dysfunction due to a dysregulated host response to infection.” It would be expected that many with COVID-19 associated mortality would meet that definition on the basis of respiratory failure. However, the striking difference between the physiology of bacterial sepsis and severe COVID-19 is that cardiovascular instability is rare in COVID-19 [19]. In our modelling of individual variables of CURB-65, we found that unlike the respiratory components, blood pressure was not independently associated with adverse outcome. Similarly, confusion, often a sign of haemodynamic compromise, was less relevant to outcomes in the CURB-65 score.

Blood pressure and mental status are integral components of the qSOFA score and in other contexts contribute to its ability to prognosticate, thus the poor performance of qSOFA is explained by the limited effect of COVID-19 on these physiological parameters. These findings suggest COVID-19 associated mortality may be mediated by different mechanisms than conventional bacterial sepsis. An example of this may be the profound endothelial injury and abundant microthrombi identified in a recent post-mortem study [22].

Given the limited performance of the previously validated and widely used scores seen here, we explored whether performance could be improved by deriving new models. Using multiple logistic regression, we derived a set of variables, CUCA-FSR and CUCA-SF, in an attempt to predict 30-day and 72-hour mortality respectively. In keeping with the findings described above, markers of cardiovascular compromise were not independently associated with poorer outcomes, with markers of respiratory function, age and frailty appearing more relevant. CUCA-SF and, to a lesser extent, CUCA-FSR both improved prognostic performance across a range of measures compared to the pre-existing scores. However, despite deriving the score *de novo* from all variables available, CUCA-SF only identified a very small proportion of admissions as ‘low risk’ and still had a residual mortality of 4% in that group. Consequently, it is unlikely to be useful for clinical decision making. An explanation for the sub-optimal outcome of this approach may lie in limitations of the data collected, as although we present a comprehensive, prospectively collected dataset from multiple centres, collection of data was pragmatic and streamlined such that only variables included in common risk scores were collected. It did not include assessment of detailed patient demographics or comorbidities, instead focusing on clinical measurements normally taken at presentation to hospital. Characteristics such as obesity, ethnicity and comorbidities are reported to be relevant to COVID-19 outcomes but are not included here [23,24]. The ISARIC4C consortium has derived a risk calculator from their large inpatient dataset using baseline patient characteristics; it may be that a combination of clinical parameters and patient characteristics is more informative than either in isolation [25].

Some limitations must be addressed. Firstly, we only included a two-week period, and it is possible demographics and outcomes may change across the course of the COVID-19 pandemic [26]. Reassuringly, the characteristics and outcomes in the study population seen here are in keeping with those reported by the ISARIC study, one of the largest studies in this setting to date. For example the median age here was 70 years compared with 72 years in the ISARIC study, 61% were male here compared with 59.9% in ISARIC, 17.1% here were admitted to critical care compared to 17% in ISARIC, and we observed 34% 30-day mortality which is comparable to the hospital case fatality rate of 33.6% reported by ISARIC [27,28]).

A further limitation here is the inclusion only of those people admitted to hospital, thus excluding those well enough to be discharged from the emergency department. As a consequence, the observed risk presented among ‘low-risk’ categories seen here may, in theory, be inflated; for example, it is possible only the sickest CURB-65 score 0-1 patients were admitted. However, the main derivation dataset for CURB-65 score only included patients admitted to hospital, replicating the methods here [5]. Similarly, a large validation study that included both admissions those discharged directly from the emergency department found 30-day mortality in those admitted to hospital with a ‘low-risk’ CURB-65 score remained low at 0.0-1.6% [16]. This supports the conclusion that the observed high mortality among low CURB-65 scores in this study was due to SARS-CoV-2 virulence rather than study design. Conversely, some patients presented to the emergency department in a moribund state and did not survive long enough for a viral swab to be taken. Such patients were not included in our study and generalisability to that small subset of patients may be limited. We defined ‘early mortality’ as death occurring within 72h of admission in order to capture patients who deteriorated quickly, but within a timeframe that would allow them to have a known SARS-CoV-2 status and be identified by our investigators. This 72h timepoint differs from the 24h timepoint used in the NEWS2 score’s validation studies but, given the above constraints, was considered a pragmatic approach for our analysis.

The strengths of this study lie in the prospective collection of data on consecutive admissions from multiple regional hospitals with rigorous assessment of the performance each score. These readily available data were compiled from real-world clinical assessment, and outcomes followed usual clinical care. We also demonstrate the hitherto underappreciated potential of highly trained and motivated speciality trainees and their ability to coordinate and collaborate for research.

## Conclusion

CURB-65, NEWS2 and qSOFA underestimate 30-day mortality among patients admitted to hospital with COVID-19. CURB-65 and NEWS2 were slightly better at predicting early mortality. However, our data suggest CURB-65 should not be used to prognosticate in the setting of COVID-19 pneumonia since low CURB-65 scores were associated with high mortality rates. We provide a set of clinical parameters which appear relevant to outcomes in COVID-19 and may provide a basis for combination with patient characteristics in ongoing efforts to improve prognostication in COVID-19.

## Data Availability

Data are available upon reasonable request.

## Contributions

Conception and design: FF, PB, DW

Clinical data collection: FF, PB, NW-CORR collaborators (listed above)

Analysis and interpretation: KT, FF, PB, DW

Manuscript preparation: FF, PB, DW, with review by KT

Clinical governance and academic supervision: DW

## Acknowledgements

We are grateful to Prof Marta Garcí a-Fiñ ana for statistical advice. Dr Abdul Ashish and colleagues at NIHR CRN Greater Manchester have kindly supported the NW-CORR trainee collaborative. Jake Gannon was instrumental in setting up our REDCap data collection tool.

## Supplementary data and figures

### List of participating organisations

Blackpool Teaching Hospitals NHS Foundation Trust, Blackpool, UK

Countess of Chester Hospital NHS Foundation Trust, Chester, UK

Liverpool University Hospitals NHS Foundation Trust, Liverpool, UK

Manchester University NHS Foundation Trust, Manchester, UK

Mid Cheshire Hospitals NHS Foundation Trust, Leighton, UK

Lancashire Teaching Hospitals NHS Foundation Trust, Preston, UK

Southport and Ormskirk Hospital NHS Trust, Southport, UK

Liverpool Heart & Chest Hospital NHS Foundation Trust, Liverpool, UK*

*=No acute admissions, outcomes recorded from critical care transfers only.

**Table S1:**
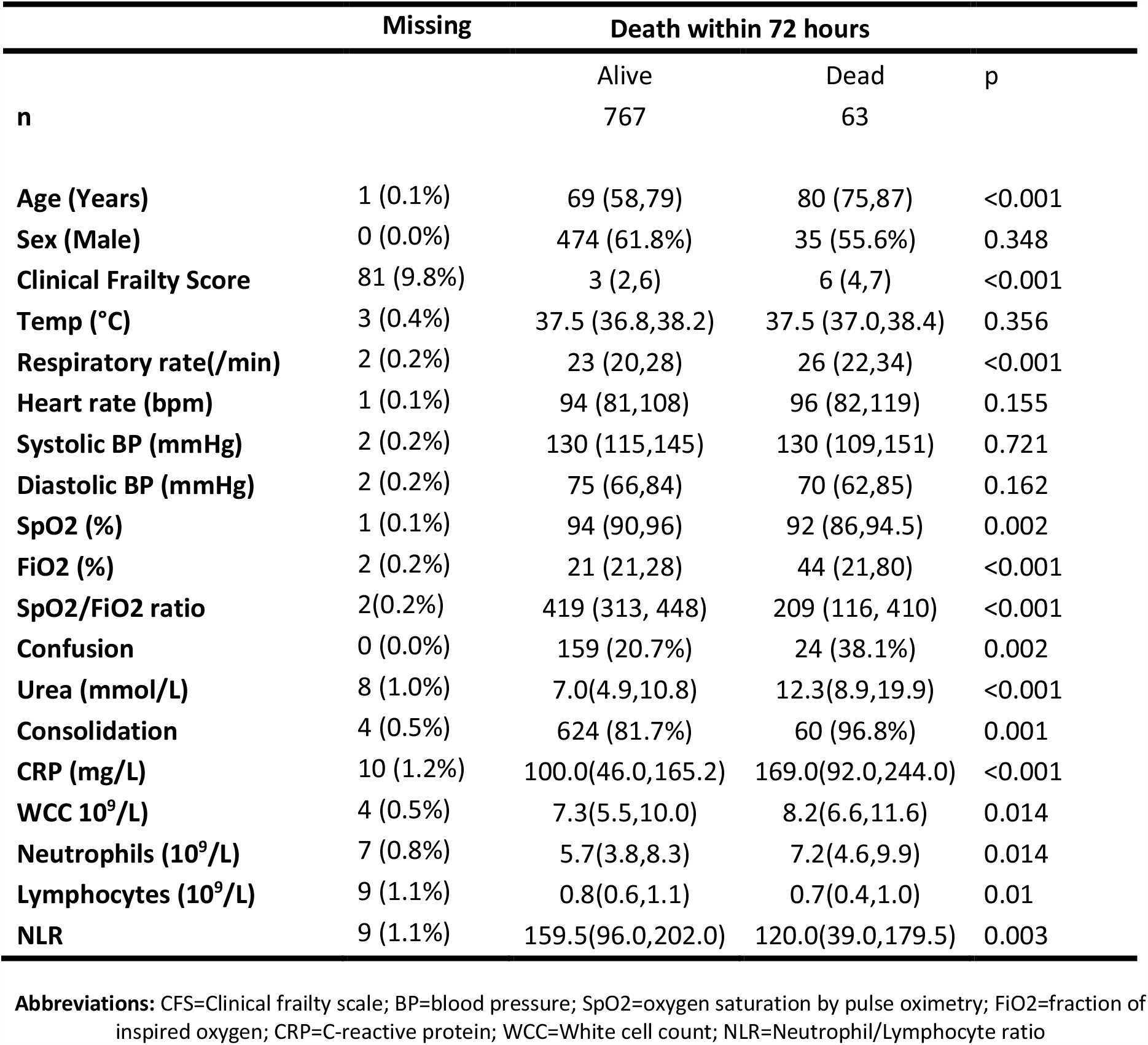
Demographics and clinical characteristics at presentation with comparison by 72-hour outcome

**Table S2:**
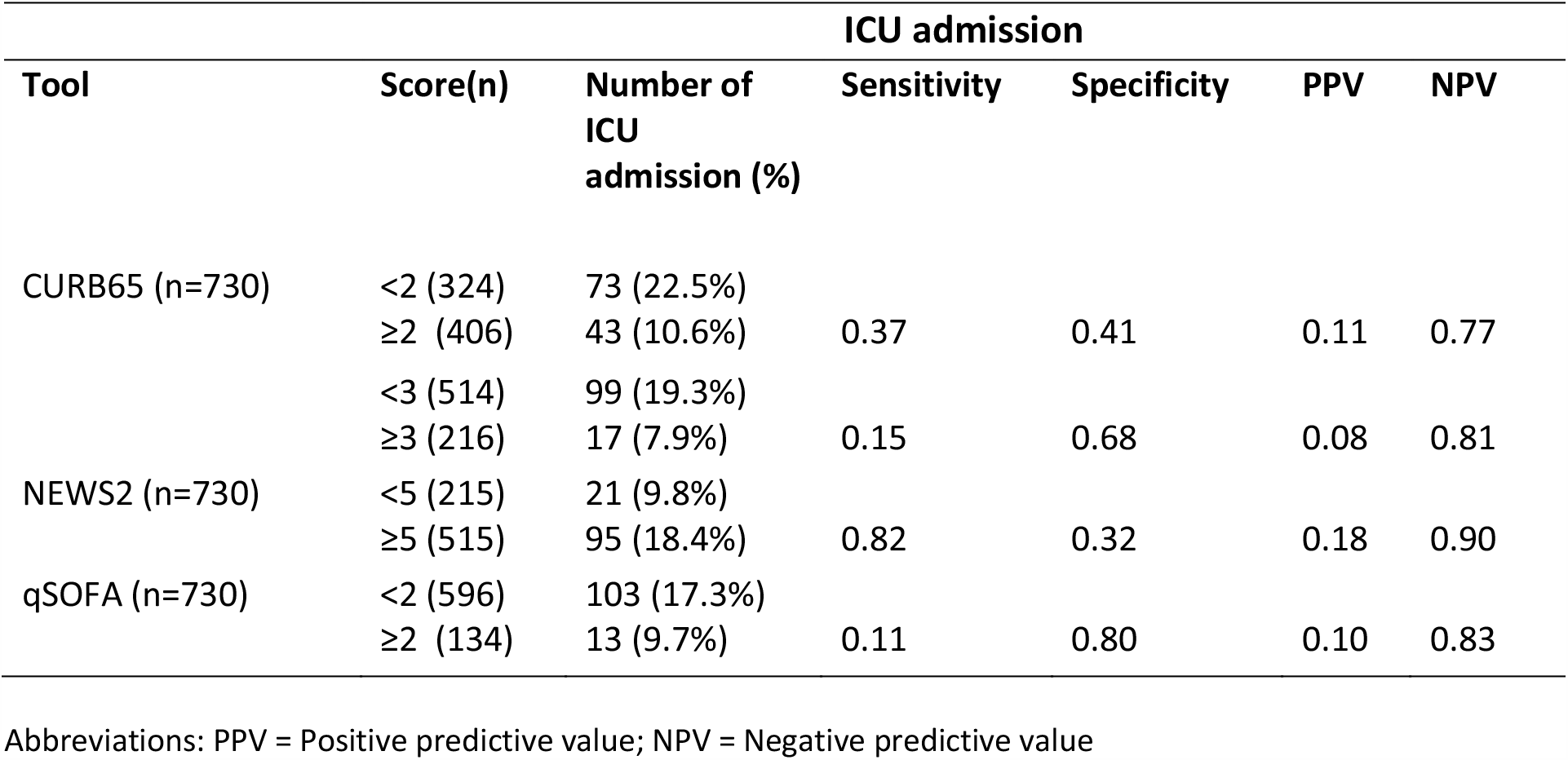
Score performance for ICU admission as outcome (sensitivity, specificity, PPV, NPV)

**Table S3:** Logistic regression models for 30-day mortality fitted using the individual constituents of each score.

| Model | Variables | OR (95%CI) | p-value |
| --- | --- | --- | --- |
| **CURB-65**<br>**(n=730)** | Confusion (Yes) | 1.29 (0.87 - 1.91) | 0.204 |
|  | Urea (>7mmol/L) | 2.90 (2.01 - 4.20) | <0.001* |
|  | Respiratory Rate (≥30) | 2.73 (1.82 - 4.08) | <0.001* |
|  | BP (SBP<90 or DBP≤60) | 1.33 (0.85 - 2.06) | 0.210 |
|  | Age (≥ 65) | 2.75 (1.86 - 4.07) | <0.001* |
| **NEWS2**<br>**(n=730)** | Respiratory Rate |  |  |
|  | 2 points | 1.29 (0.84 – 1.97) | 0.245 |
|  | 3 points | 1.90 (1.25 – 2.90) | 0.003* |
|  | SpO2 |  |  |
|  | 1 point | 0.81 (0.50 – 1.30) | 0.379 |
|  | 2 points | 1.59 (0.97 – 2.60) | 0.064 |
|  | 3 points | 1.47 (0.99 – 2.20) | 0.058 |
|  | FiO2 |  |  |
|  | 2 point | 2.00 (1.43 – 2.80) | <0.001* |
|  | Temperature |  |  |
|  | 1 point | 1.43 (0.99 – 2.06) | 0.059 |
|  | 2 points | 1.68 (0.90 – 3.14) | 0.102 |
|  | 3 points | 1.33 (0.32 – 5.42) | 0.694 |
|  | Systolic BP |  |  |
|  | 1 point | 1.19 (0.68 – 2.06) | 0.541 |
|  | 2 points | 2.06 (1.04 – 4.06) | 0.038* |
|  | 3 points | 1.23 (0.56 – 2.70) | 0.606 |
|  | Heart rate |  |  |
|  | 1 point | 0.65 (0.45 – 0.95) | 0.026* |
|  | 2 points | 0.78 (0.49 – 1.25) | 0.303 |
|  | 3 points | 0.84 (0.39 – 1.79) | 0.651 |
|  | Confusion (Yes) | 2.35 (1.63 – 3.40) | <0.001* |
| **qSOFA**<br>**(n=730)** | Respiratory Rate (≥ 22) | 1.85 (1.33 - 2.58) | <0.001* |
|  | Systolic BP (≤100) | 1.74 (1.04 - 2.91) | 0.035* |
|  | Altered mental status (Yes) | 2.37 (1.66 - 3.38) | <0.001* |

**Table S4:** Score performance for ICU admission as outcome (sensitivity, specificity, PPV, NPV)

| Tool | Score(n) | Number of ICU admission (%) | ICU admission |  |  |  |
| --- | --- | --- | --- | --- | --- | --- |
|  |  |  | Sensitivity | Specificity | PPV | NPV |
| CAS-FC<sup>1</sup> (n=730) | <1(381) | 11 (2.9%) |  |  |  |  |
|  | ≥1 (349) | 105(30.1%) | 0.91 | 0.60 | 0.30 | 0.97 |
|  | <2 (589) | 47 (8.0%) |  |  |  |  |
|  | ≥2 (141) | 69 (48.9%) | 0.59 | 0.88 | 0.49 | 0.92 |

Abbreviations: PPV = Positive predictive value; NPV = Negative predictive value

## Notes

### Competing Interest Statement

The authors have declared no competing interest.

### Clinical Trial

Institutional approval for data collection and anonymised data collation was obtained from all sites and were a combination of Caldicott guardian and research/audit committee approvals. No approach to the patient was made and only fully anonymised routinely available clinical information was collated; on this basis consent was not required under guidance from the NHS Human Research
Authority.

### Funding Statement

No external funding was received.

### Author Declarations

Institutional approval for data collection and anonymised data collation was obtained from all sites and were a combination of Caldicott guardian and research/audit committee approvals.

